# Involvement of the Posterior Visual Pathway Correlates with Higher-Order Visual Impairment in Childhood Stroke Patients detected by Virtual Reality/Eye Tracking Paradigm

**DOI:** 10.1101/2024.11.03.24316675

**Authors:** Emily Da Cruz, Diana Tambala, Anna Lynch, Claire Manley, Melissa Bambery, Daniel Kelly, Carrie Chui, Kenda Alhadid, Alyssa W. Sullivan, Julie Grieco, Benjamin Ondeck, Arne Lauer, Lotfi B. Merabet, Patricia L. Musolino

**Author notes:** <u>Corresponding Author:</u> Patricia L. Musolino, MD, PhD, Co-Director Pediatric Stroke and Cerebrovascular Program, Associate Professor of Neurology Harvard Medical School, 185 Cambridge St, Suite 5248, Boston, MA 02114. The authors share first authorship (Emily Da Cruz, Diana Tambala, Anna Lynch). The corresponding authors supervised the project equally (Arne Lauer, Lotfi B. Merabet, Patricia L. Musolino).

## Abstract

**Background/Objective:** Cerebral injury due to stroke in childhood increases the risk of higher-order visual processing (HOVP) deficits, like cerebral visual impairment (CVI), which can lead to severe behavioral and learning disabilities if left untreated. Using a virtual reality-based search task and structural Magnetic Resonance Imaging analysis, we assess the extent of functional vision deficits in childhood stroke patients and potential anatomical correlates.

**Methods:** 20 childhood stroke patients and 38 healthy controls completed a dynamic visual search task using a virtual reality/eye-tracking (VR/ET) paradigm to quantify functional vision abilities between 2021 to 2024 (average 7.34 years after stroke). Virtual reality assessment measures, stroke imaging characteristics (visual pathway involvement) and neuropsychological outcomes were analyzed between cohorts using statistical comparison methods and linear regression model.

**Results:** All childhood stroke patients could complete the VR/ET task, whose metrics were associated with neuropsychological testing measures of visual attention and processing speed, as demonstrated by success rates and task compliance in equal measure to controls. However, less accurate search and slower fixation rates together with less sensitivity to changes in task load and greater impairment in initiating a response to a target were observed in our patient cohort. On MRI lesion analysis injury involving the posterior visual pathways, specifically the optic radiations, inferior longitudinal fasciculus, or superior longitudinal fasciculus, correlated with slower reaction time to fixation on a target when controlling for age at time of VR testing.

**Conclusions:** Bedside VR/ET assessment in children affected by stroke can detect signs of HOVP deficits as confirmed by neuropsychological testing. Imaging demonstrating involvement of the posterior visual pathway at the time of diagnosis is strongly correlated with development of impaired visual tracking abilities later in life. While detection of HOVP deficits relies on current standard clinical and neuropsychological evaluations between 3 to 6 years of age, our study demonstrates that injury pattern on imaging at stroke onset can help identify children at risk of HOVP deficits. This may enable early monitoring and timely accommodations facilitating functional vision development, critical to learning and skill acquisition.

## Introduction

Stroke in childhood occurring during the perinatal period (28 weeks of gestation to 28 days of life) or pediatric (29 days to 18 years of age) is one of the leading causes of childhood disability^1,2^. More than 50% of perinatal and pediatric stroke survivors experience a cognitive/sensory disorder impacting their quality of life, making early detection critical to determining early interventions that could improve outcomes^3-7^. While motor deficits become apparent early on, most behavioral or cognitive deficits only become apparent as developmental stages are attained and vulnerabilities become outwardly observable, thus delaying implementation of rehabilitation interventions^8-10^. Critical to the development of higher cognitive functions are higher-order visual processing (HOVP) systems, defined as the cognitive abilities of integrating visual input for goal-oriented tasks^11,12^. Visual field loss^13^, oculomotor dysfunction, and visual perceptual impairments^13-15^ have been extensively studied in adult stroke survivors. Studies assessing HOVP found that up to 30% of survivors suffer from visual inattention, impaired functional vision (visual task-related ability) and/or difficulty with adaptation to changes in environmental stimuli^7,16^. Presence of HOVP impairments was determined by measuring the ability to modulate responses to visuospatial cues^17^, reaction time and accuracy in goal-oriented visuospatial tasks^18^, or assessing reading difficulties due to eye movement abnormalities^19^. Recently, virtual reality (VR) combined with eye tracking (ET) paradigms have been used to assess HOVP in a naturalistic environment while eliminating the reliance on the individual’s other abilities that stroke can affect, including verbal and motor abilities^20-23^.

The current NIH-endorsed working definition of cerebral visual impairment (CVI) describes it as an HOVP disorder characterized by a range of visual impairments resulting from early damage and/or maldevelopment of visual pathways and processing centers of the brain^11,24-26^. While visual acuity is regularly assessed in children, functional vision is not typically included in early ophthalmological or neurological assessments in perinatal and pediatric stroke survivors. Typically diagnosed by optometrists or ophthalmologists, CVI requires a multidisciplinary approach with considerations from ophthalmologists, neurologists, psychologists, and other specialists to rule out explanations provided by ocular abnormalities and disorders of behavior, learning and/or social communication while also confirming a neurologic condition affecting the visual pathway and deficits in HOVP^27^. Of interest is the neuropsychological evaluation, which provides a quantitative metric in assessing visuospatial perception and constructional abilities, as well as visual attention, learning, memory capacities, and potential comorbid behavioral disorders^28^. Early detection of CVI in children is critical as early implementation of interventions can maximize rehabilitative neuroplasticity ^7,29,30^. Implementing individualized education program based vision and orientation/mobility services, as well as accommodations for vision are often indicated with modifications such as: elimination of eliminate environmental clutter^31^, mask surrounding text while reading, and encourage educators to wear distinct, bright clothing to aid in identification^32^, is recommended for children with CVI. If left unaccommodated for, functional vision deficits can create additional barriers to success and impede children’s access to education and rehabilitative therapies, development, and functional independence^33^.

In this study, we sought to quantify childhood stroke survivors’ HOVP impairment as compared to healthy controls using a VR/ET paradigm and correlate their VR/ET performance with stroke lesion characteristics on brain imaging to identify anatomical correlates to their HOVP abilities.

## Methods

### Participants

The study included two cohorts of participants: childhood stroke survivors (perinatal and pediatric stroke) and controls (neurotypical development with normal visual acuity and no CVI). Participants were recruited and enrolled from June 2021-January 2024. Members of the stroke cohort were recruited from the MGH (Massachusetts General Hospital) Multidisciplinary Stroke Clinic in Boston, MA, during clinic visits. Members of the control cohort were recruited from the Boston area. Details of control participant recruitment are described in Manley et al. 2022^21^. They completed the VR assessment following their consent.

The study was approved by the local Institutional Review Board, Mass General Brigham Human Research Committee (MGBHRC) prior to participant enrollment and included both a retrospective medical records protocol (2015P002030) and a prospective protocol (2019P000070). For the retrospective protocol, participant consent was waived by the MGBHRC. For the prospective protocol, written informed consent was obtained from all participants (or guardians of participants). This study was carried out in accordance with the Code of Ethics of the World Medical Association (Declaration of Helsinki) for experiments involving humans.

### VR Testing

The virtual hallway environment was developed using the Unity 3D game engine version 5.6 (Unity Technologies, San Francisco, CA, USA). Participants sat a viewing distance of 50-60cm (about 1.97 ft) away from a 27” laptop monitor, with the eye tracker unit (Tobii 4C; 90 Hz sampling frequency. Tobii Technology AB, Stockholm, Sweden) mounted on the lower portion of the monitor. The virtual hallway stimulus subtended 58 × 32 degrees of visual angle. Participants were able to move their heads freely but reminded to keep their gaze on the screen throughout testing. Three-dimensional object models were created using Blender modeling software (Blender Foundation), and 3D human models were created in Adobe Fuse CC and rigged for animation in Adobe Mixamo (Adobe Systems Inc., San Jose, CA, USA)^22^.

The virtual hallway is a dynamic search task in which participants are instructed to scan a screen, displaying the hallway of a fictitious school with a crowd of animated characters walking toward the observer (Fig. 1). From a fixed, first-person perspective, participants are instructed to locate and pursue the principal each time she appears on the screen from one of the eight entrances until she exits the scene. The interval between a target disappearing and reappearing in the hallway from trial to trial varied between 5 and 15 sec. The duration of the target’s visibility was determined by its starting point and path length. This varied between 5 and 17 sec for the closest and furthest starting points, respectively. Crowd density (the number of characters on screen at a given time) was manipulated by three categories: low (average of 5 ± 5 people), medium (10 ± 5), and high (15 ± 5), with each level of crowd density presented equally and in a pseudo-random fashion. Each trial run lasted about 3.5 minutes, and participants completed 2 trial runs with a brief break in between. For further details regarding the design of the visual stimulus and task, see Bennett et. al., 2022^21^.

**Figure 1.**
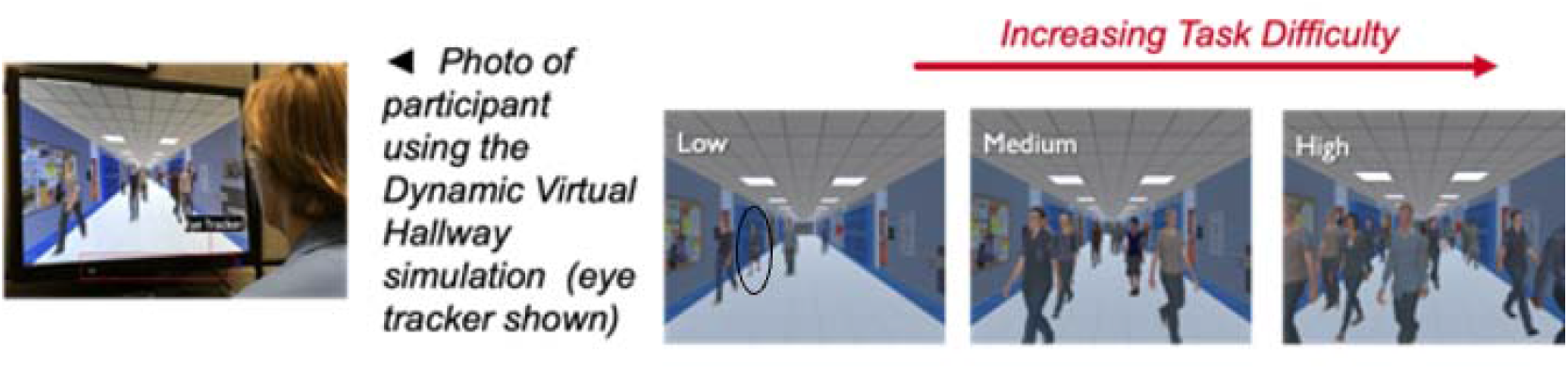
Dynamic virtual hallway VR assessment setup. The dynamic search task tracks participants’ eye movements as they are tasked with searching and pursuing a character (denoted as the principal, circled) as various “distractor” characters walk in and out of the hallway on screen. Crowd density fluctuates throughout the trial from Low (5 ± 5 people), Medium (10 ± 5 people), and High (15 ± 5 people) task difficulty. The calibrated eye tracker collects visuomotor data, then processed as the outcome metrics of the task.

### Outcome Measures

Eye gaze behavior was processed using an in-house Matlab script (MathWorks, Natick, MA, USA). Metrics used for analysis were success rate, reaction time, gaze error, search area, and off-screen percentage. Success rate is the percentage representing how many times the participant located and fixated on the target (the principal) in each trial. Reaction time is defined as the first moment gaze within the target’s outer contour and remains fixated for the presentation. Gaze error is a measure of fixation accuracy assessed as the distance between the center of the target and the participant’s gaze position. Search area represents the visual search area explored and serves as an index for search precision; it is measured by an ellipse-shaped 95% confidence interval fitted to the captured eye-tracking data. Off-screen percentage measures task compliance, assessing the proportion of gaze points falling outside the bounds of the screen on each trial.

### Retrospective Medical Record Review

Childhood stroke participants’ medical records and brain multimodal magnetic resonance imaging (MRI) were reviewed to characterize and segment the cohort based on different stroke characteristics. A neuroradiologist (A.L.) blinded to clinical information and the VR/ET task results assessed the MRI closest to the incident of stroke. Stroke injury was assessed on multi- modal axial and/or coronal planes and assigned a binary categorization of “yes” or “no” to determine injury involving structures comprising the visual pathway. This determination was based on whether chronic or acute stroke lesions involved the optic nerve, optic tract, LGN,OR, occipital lobe, SLF, ILF. Stroke injury was further categorized by visual pathway involvement, categorizing higher-order involvement (parieto-occipital or occipito-temporal lobe) and lower-order involvement (occipital pole). To further characterize behavioral abilities, data regarding attention-deficit/hyperactivity disorder (ADHD), and neuropsychological evaluations were extracted after consultation with a psychology fellow (A.S) and neuropsychologist (J.G.). Scaled scores (mean = 10, sd =3) from two subtests from the Wechsler Intelligence Scale for Children - 5^th^ edition (WISC-V)^34^ (Coding, Symbol Search) were compiled for analysis based on anticipated concurrent validity with HOVP and reaction times.

### Statistical Analysis

Statistical analyses were carried out using GraphPad Prism version 10.0.2(171) (Boston, MA, USA). The Kolmogorov-Smirnov test was used to test for normality. For each behavioral outcome, an unpaired t-test, ANOVA, Mann-Whitney U, or Kruskal-Wallis test (dependent on normality) was conducted with cohorts as a between-subjects factor and task difficulty (i.e., low, medium, and high) as a within-subjects factor. For ANOVA and Kruskal-Wallis analyses, Dunn’s test to correct for multiple comparisons was carried out on mean differences of behavioral outcomes to explore the directionality of the observed effects. Between-group comparisons were planned between cohorts. Within-group comparisons were planned for reaction time based on task difficulty in childhood stroke and control cohorts. A series of within-group analyses were conducted in the childhood stroke cohort and can be found in the supplemental text. Multivariable linear regression models were created to determine if VR task reaction time correlated with 1) age at VR testing, 2) neuropsychology standard scores when controlling for PVP injury (binary yes/no), and 3) stroke injury involvement of anatomical structures comprising the visual pathway when controlling for age at VR testing. Results were considered significant if p<0.05.

Based on earlier work in this group^21,22^, we have found that reaction time is the most sensitive and has the largest effect of the VR metrics. Therefore, we will use this as the primary outcome metric when correlating VR/ET task performance with variables including age at time of VR testing, neuropsychology standard scores, and injury involvement of visual pathway anatomical structures.

### Data Availability

Anonymized data not published within this article will be made available by request from any qualified investigator.

## Results

### Childhood stroke cohort characteristics and feasibility of VR task

We studied participants in two cohorts: 1) a confirmed pediatric or perinatal stroke diagnosis as confirmed by MRI and no formal diagnosis of CVI (n=20, mean age: 9.9 years, range: 6-15 years; median: 9; inter-quartile range (IQR): 4) and 2) controls with neurotypical development and no history of ophthalmic (e.g., strabismus, amblyopia) or neurodevelopmental conditions (n=38, mean age: 20.9 years, range: 11-33 years; median: 20; IQR: 4) (Table 1). All participants had normal or corrected-to-normal visual acuity and no visual field deficits. On average, patients had an MRI 71.9 days after the stroke. Of the childhood stroke cohort, 11 patients have experienced perinatal stroke, and 9 pediatric stroke. VR/ET testing happened 7.34 years after the stroke. Majority of stroke lesions were ischemic (65%), involved the MCA vascular territory (60%), and affected males (70%). Neuropsychological evaluations were available in 17 stroke patients, between the ages of 5 and 15 years, and the evaluation closest to the time of VR testing was analyzed. Further stroke cohort characteristics are detailed in Table 2.

**Table 1.** Cohort Demographics.

| Table 1. Cohort Demographics |  |  |
| --- | --- | --- |
|  | Childhood Stroke | Controls |
| Age at VR/ET testing, mean (range) | 9.9 years (6-15 years) | 20.9 years (11-33 years) |
| *Sex, n (%)* |  |  |
| Male | 14 (70%) | 12 (31.6%) |
| Female | 6 (30%) | 26 (68.4%) |

**Table 2.** Childhood Stroke Cohort (n=20) Characteristics.

| **Table 2. Childhood Stroke Cohort (n=20) Characteristics** |  |
| --- | --- |
|  | ***n (%)*** |
| *Stroke Timing* |  |
| Perinatal Stroke (28 weeks of gestation to 28 days of life) | 11 (55%) |
| Pediatric Stroke (29 days to 18 years) | 9 (45%) |
| *Stroke Type* |  |
| Ischemic | 14 (65%) |
| Hemorrhagic | 4 (25%) |
| Venous Infarction | 2 (10%) |
| *Stroke Territory* |  |
| ACA | 2 (10%) |
| MCA | 12 (60%) |
| PCA | 2 (10%) |
| Cerebellar | 3 (15%) |
| Others | 4 (20%) |
| *Posterior Visual Pathway Involvement* |  |
| Left | 7 (35%) |
| Right | 2 (10%) |
| Bilateral | 1 (5%) |
| *Posterior Visual Pathway Injury Type* |  |
| Acute | 8 (40%) |
| Subacute | 2 (10%) |
| *Visual Pathway Involvement* |  |
| Higher-Order Visual Involvement | 4 (20%) |
| Lower-Order Involvement | 5 (25%) |
| No Visual Pathway Involvement | 11 (55%) |
| *Stroke Laterality* |  |
| Left Hemisphere | 8 (40%) |
| Right Hemisphere | 5 (25%) |
| Bilateral | 3 (15%) |
| *Comorbidities* |  |
| ADHD | 7 (35%) |
| Cerebral Palsy | 11 (55%) |
| *Neuropsychological Testing* |  |
| Wechsler Intelligence Scale for Children-5th ed. (WISC-V) | 16 (80%) |
| *Interval between Neuropsychological and VR/ET Testing* |  |
| Interval in years, mean (range) | 1.97 years (0.13-5.81 years) |

No significant differences in VR/ET metrics were observed between perinatal and pediatric stroke cohorts (Mann-Whitney *U* tests: success rate: p=0.334, reaction time: p=0.992; gaze error: p=0.552; search area: p=>0.999; off-screen percentage: p=0.807) (Supp. Fig. 1). Both cohorts were therefore combined as a single childhood stroke cohort for subsequent analysis. This cohort’s ability to perform VR/ET tasks was confirmed similar to controls success rate (stroke: 94.72±6.62; controls: 97.66±3.03; Mann-Whitney *U*: p=0.162) and off-screen percentage values (stroke: 0.36±0.42; controls: 0.38±0.29; Mann-Whitney *U*: p=0.285) (Fig. 2A).

**Figure 2.**
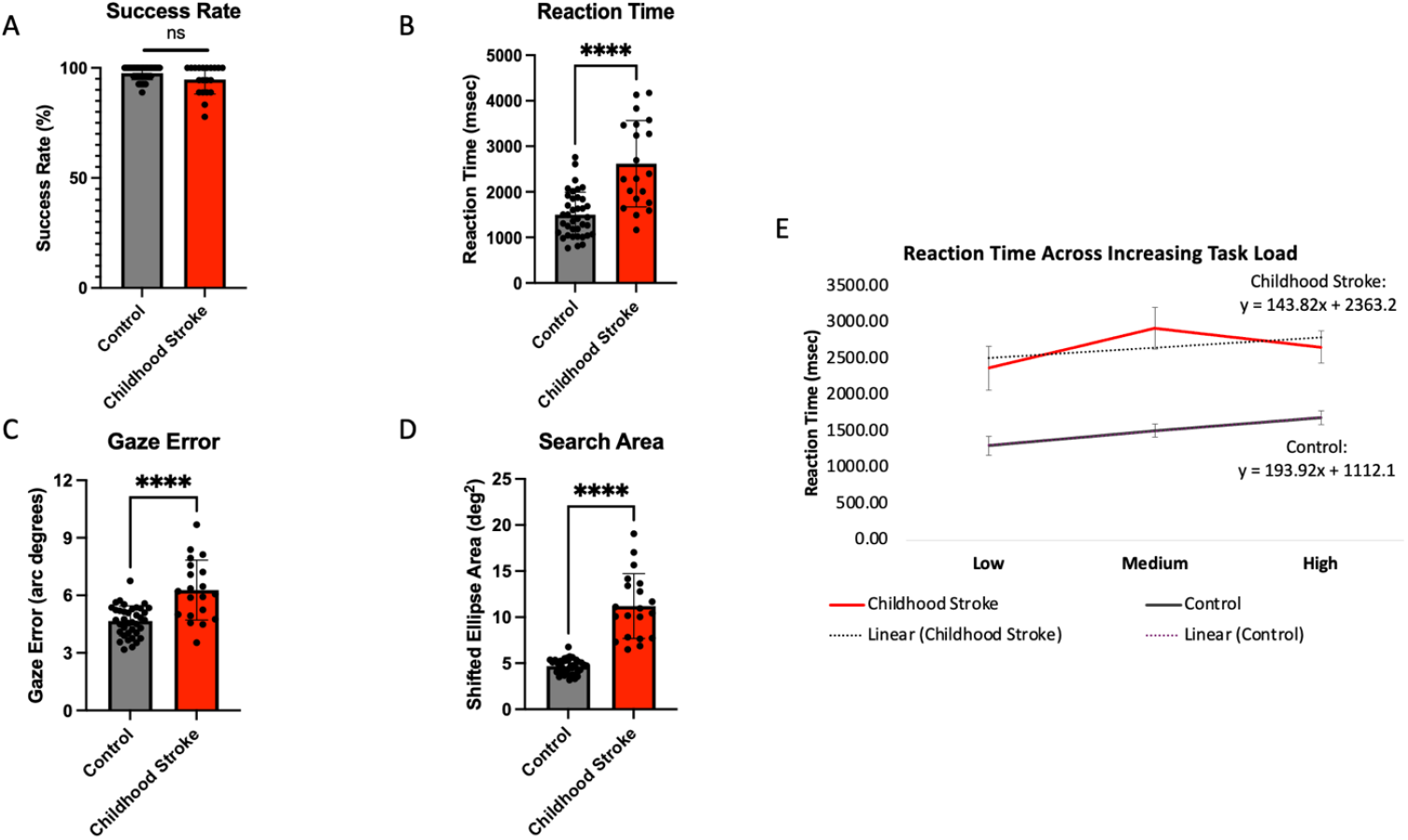
Oculomotor metrics of the childhood stroke and control cohorts. Despite similar task success compared to controls **(A)**, stroke patients showed significant differences in **(B)** reaction time (p<0.0001), **(C)** gaze error (p<0.0001), and **(D)** search area (p<0.0001). **(E)** When assessing changes to reaction time across the varying task load, control participants have a steeper change in reaction time than childhood stroke patients, as shown by slope values of best-fit lines. Further, the childhood stroke cohort has a higher initial visual orienting response, as calculated by the extrapolated y-intercept, than control participants.

Participants’ age showed no effect on VR performance assessed by reaction time, despite ranges in age both within and across the childhood stroke and control cohorts [stroke cohort linear regression: p=0.7882 and R^2^ = 0.004355; control cohort linear regression p=0.7339 and R^2^ = 0.003344].

When assessing how VR/ET performance correlated with patients’ neuropsychological testing performance on two tasks evaluating visual attention and scanning efficiency, we find that when controlling for stroke injury involving the PVP, reaction time on the VR/ET paradigm correlates with the scaled score for the symbol search subtest of the processing speed domain in WISC-V (p=0.041; R^2^ = 0.3820).

### Childhood stroke patients show significant deficits in VR search task and decreased adaptive capacity to increased complexity of the visual environment

Childhood stroke patients had significantly higher reaction time to fixation (mean±SD: stroke: 2617.76±948.72; controls: 1499.82±492.21; t-test: p<0.0001), a larger gaze error (stroke: 6.27± 1.56; controls: 4.67±0.79; t-test: p<0.0001;), and a larger search area compared to controls (stroke:11.20±3.51; controls: 6.97±2.17; t-test p<0.0001) (Fig. 2B-D). Reaction time was assessed across the “low,” “medium,” and “high” task difficulty levels in both cohorts. Visual search was tested at increasing visual environment complexity from low to medium to high levels. While there was no significant difference across task complexity levels for reaction time in the childhood stroke cohort (low: 2371.81±1347.31; medium: 2921.16±1293.13; high: 2659.44±1000.67; Kruskal-Wallis: p=0.264), the control cohort exhibited significant differences (low: 1301.09±797.54; medium: 1509.96±582.61; high: 1688.93±581.55; Kruskal-Wallis: p=0.002). This resulted in a less steep slope for reaction time in the stroke cohort (slope=143.8 vs 193.92 of controls) denoting an impaired visual processing adaptive capacity. Additionally, the initial visual orienting response, or the reaction time to identify the target in the absence of distractors, as defined by the extrapolated y-intercept value of the reaction time at different levels of task complexity, was higher (y-intercept=2363.2) in childhood stroke patients than controls (y-intercept=1112.1) (Fig. 2E).

### Posterior visual pathway involvement of stroke predicts higher-order visual processing deficits

In analyzing the stroke injury involvement in the anatomical structures in the visual pathway (optic nerve, optic tract, LGN, occipital lobe, OR, SLF, and ILF), we found that stroke patients with lesions involving the posterior structures of the visual pathway on MRI correlated with longer reaction times on the VR assessment after controlling for age at VR assessment. Specifically, injury involving the OR (n=7), SLF (n=9), or ILF (n=6) all correlated with increased reaction time when controlling for age at VR assessment (p= 0.0327, 0.0311, and 0.0031, respectively) (Figs. 3 and 4). However, stroke injuries involving the optic nerve, optic tract, LGN, or occipital lobe structures of the pathway did not correlate with reaction time when controlling for age at VR assessment. Further, laterality of visual pathway injury and time of stroke injury diagnosis (acute or subacute) did not show significant differences in reaction time (laterality: p=0.127; acute vs. subacute imaging: p=0.333). When assessing differences in reaction time based on involvement of the visual pathway, those with visual pathway involvement had significantly higher reaction times than those with stroke injuries not involving the visual pathway (p=0.043). However, there was no significant difference when comparing the reaction times of those categorized as having a stroke with lower-order involvement, higher-order involvement, and no visual pathway involvement (p=0.0578).

**Figure 3.**
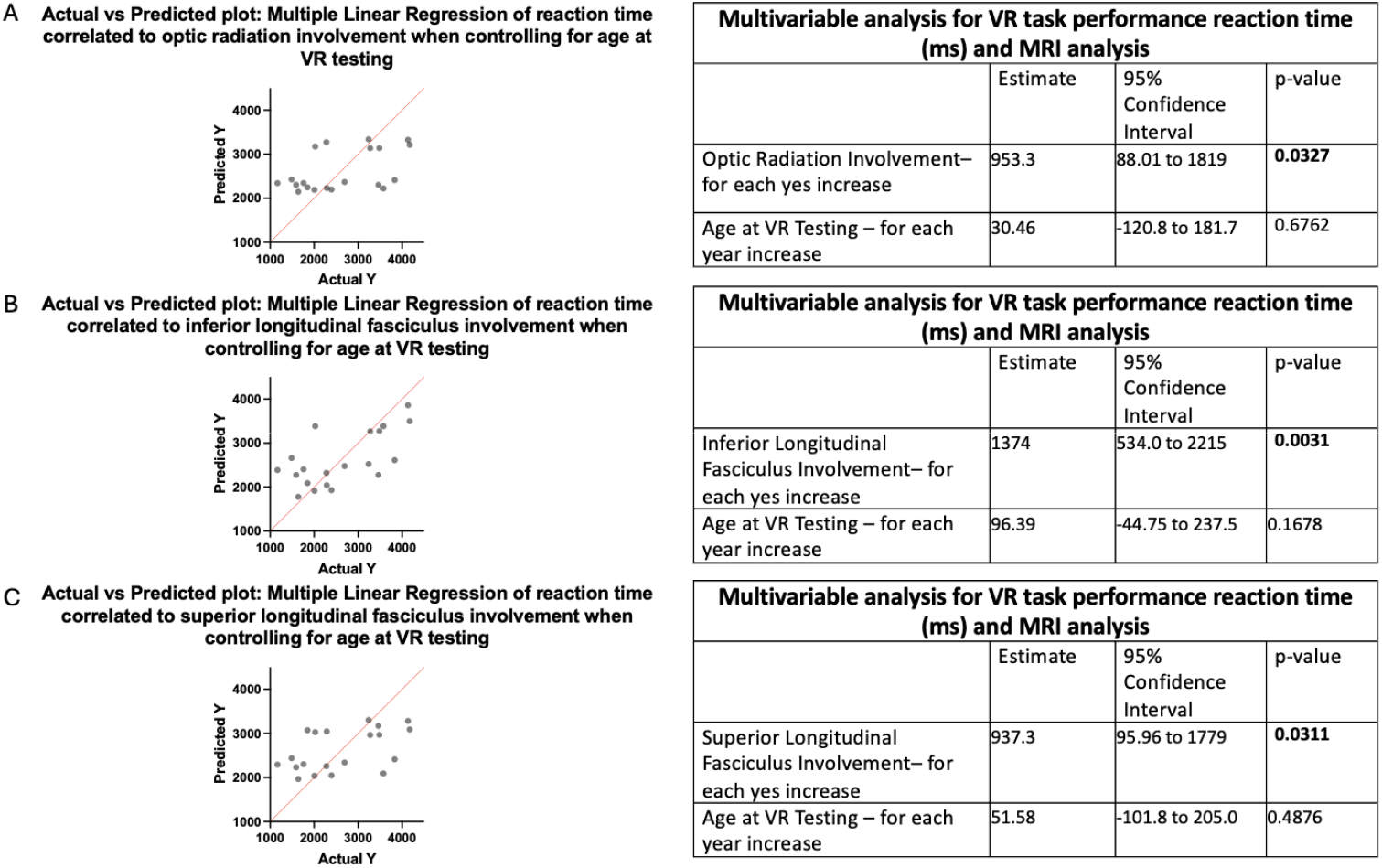
Posterior visual pathway (PVP) involvement and correlation with VR/ET in childhood stroke participants. **(A)** When controlling for age at VR testing, childhood stroke patients with stroke injury involving the OR significantly correlated with reaction time in the VR search task (p=0.0327). **(B)** When controlling for age at VR testing, childhood stroke patients with stroke injury involving the ILF significantly correlated with reaction time in the VR search task (p=0.0031). (C) When controlling for age at VR testing, childhood stroke patients with stroke injury involving the SLF significantly correlated with reaction time in the VR search task (p=0.0311).

**Figure 4.**
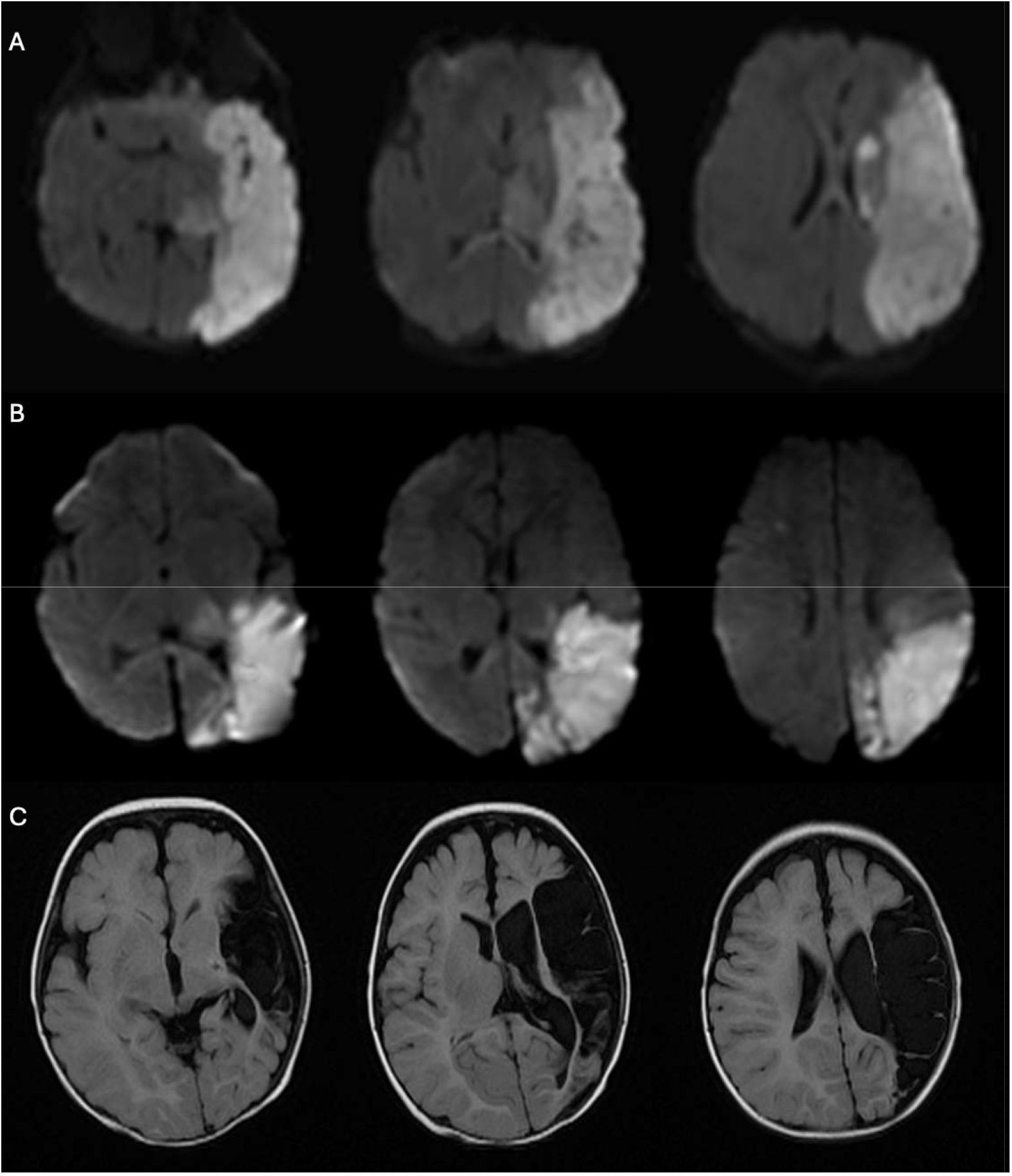
Representative MR images of posterior visual pathway (PVP) involvement. **(A, B)** Diffusion-weighted imaging (DWI) representative of a large ischemic stroke shows acute involvement of the left OR, SLF, and ILF. **(C)** FLAIR imaging representative of chronic ischemic stroke with acute involvement of left OR, SLF, and ILF.

## Discussion

In this study, we demonstrated that VR/ET is a sensitive and feasible non-invasive method to assess HOVP in a cohort of children affected by childhood stroke that reflects the quantitative assessments used in neuropsychological evaluations to help diagnose HOVP disorders. Our significant correlation between VR/ET reaction time and normative-based, standardized neuropsychological task assessing visual attention and scanning efficiency suggests that an aspect of functioning included in HOVP (visual processing speed) can be measured via the VR/ET paradigm and is associated with neuropsychological measures of a similar process. While no clinical diagnosis of HOVP disorders or CVI was present at the time of assessment, all childhood stroke patients in our study showed significantly decreased visual processing abilities compared to healthy children. Our VR/ET paradigm has the potential to screen childhood stroke participants for HOVP deficits in a naturalistic and engaging test. For the first time, we show that involvement of the PVP on brain imaging at the time of stroke diagnosis is associated with significantly increased incidence of HOVP deficits, providing an early diagnostic tool to identify at-risk patients to guide timely diagnosis and initiation of adaptive and habilitating interventions. If imaging indicates PVP involvement, specifically in the OR, SLF, or ILF, in childhood stroke injuries, providers could further examine HOVP deficits. For example, PVP involvement identified by MRI could be coupled with formal assessments of functional vision to guide accommodative practices for children’s visual processing deficits during early school years critical to learning and higher cognitive function development. In tandem with findings from neuropsychological evaluations, providers can identify the domains in which children struggle and provide individualized accommodations.

Our study is consistent with findings on visual processing impairments in adult stroke survivors, as we identify similar delays in task-related visuospatial tasks and inability to sufficiently modulate their performance to changes in environmental stimuli^17,18^. Identification of HOVP disorders and CVI in children may impact their life-long developmental potential given their increased potential for neuroplasticity. Given that focal and diffuse white matter injury, when addressed early in development and close to stroke onset, the young brain can use remapping to compensate for functional loss at the injury site^35^.

Further, our study affirms the existing literature highlighting the importance of the OR, SLF, and ILF as white matter structures crucial for relaying visual stimuli to higher cortical structures to execute HOVP abilities while filling the gap specific to childhood stroke. At the anatomical level, HOVP requires function of the visual pathway. In particular the posterior structures are responsible for relaying visual stimuli to cortical areas responsible for higher-order abilities. White matter structures such as the optic radiations (OR)^36,37^, inferior longitudinal fasciculus (ILF)^38^, and superior longitudinal fasciculus (SLF)^39,40^ are critical in the process of transmitting such information from anterior structures of the visual pathway, such as the optic nerve, optic tract, and lateral geniculate nucleus, and have been shown to be linked to visual processing abilities^37,39-41^. From the occipital lobe, there are two main streams or pathways: 1) the ventral stream to the temporal lobe, involved in object identification and recognition; and 2) the dorsal stream to the parietal lobe, involved in object’s spatial location and visual input to motor functions^42-44^. Injury to the posterior visual pathway (PVP), commonly identifiable in imaging after stroke, has been correlated with visual field deficits^45,46^, poor visual function^37,47^, and impaired visual acuity^32^.

While Berman et. al.^36^ and Bassi et. al.^37^ both demonstrate that white matter development of the OR is directly related to visual function in preterm infants susceptible to white matter injury, our findings add to the discussion by identifying a relation between this structure and visual function, as measured by reaction time, in our cohort of childhood stroke-based white matter injury. Further, our significant findings correlating SLF involvement in injury to slower reaction times on the VR/ET task reflect Kim et. al.’s^41^ findings that SLF microstructure changes in adult stroke survivors correlated with worse visual perception test performance, while also extending the conversation to childhood stroke. Existing literature demonstrating the ILF’s role in object^40^ and facial recognition^39^ contextualizes the reason we see a correlation between the VR/ET task reaction time and stroke injury involving the ILF, further emphasizing the importance of this white matter structure in functional vision.

As previously shown, special education programming can mitigate HOVP and/or CVI-related impairments that affect a child’s ability to access academic curriculum and make developmental progress. Services and accommodations, including decluttering classroom environments^31^ and additional visual cues to signal visual attention^32^ yield benefits. Similarly, orientation and mobility services can ensure safe ambulation and attention to spatial surroundings, promoting independence and inclusion. These accommodations will help stroke patients complete goal-based tasks despite their decreased adaptive capacity to increased visually complex environments as identified in the VR/ET paradigm. Recognizing ways to tailor learning environments to the needs of stroke survivors can improve their overall experience and given age-dependent plasticity, early diagnosis and intervention can improve successful remapping^35^. Therefore, utilizing accommodations recommended for CVI-diagnosed patients, such as masking texts when reading and eliminating unnecessary visual stimuli on worksheets, may improve childhood stroke survivors’ quality of life.

The limitations in our study include lacking age-matched stroke and control cohorts. However, we found that age didn’t affect success rate, reaction time, search area, or off-screen percentage metrics but did affect gaze error. A low sample size for the stroke cohort limited the power of subanalyses assessing potential differences between stroke patients with varying degrees of visual pathway involvement and further stroke characteristics, such as stroke type or vascular territory. A binary assessment of the involvement of anatomical structures in stroke lesions on MRI (present or absent) doesn’t account for the degree of injury or loss of function and limits the degree to which we can comment on anatomical involvement of stroke injury in structures. Our data is inclusive of various stroke subtypes, reflecting a clinically relevant and heterogeneous sample of childhood stroke patients. To our knowledge, this is the first study in children that characterizes visual processing deficits and their correlation to anatomical injury at time of diagnosis providing a predictive tool to guide diagnostic workup and earlier meaningful interventions.

Future work evaluating this technique in a larger cohort would validate our findings and permit further analysis of outcomes of different stroke factors and neuropsychological testing correlates. Assessment and mapping of the functional connectivity of brain regions could provide detailed insight into the correlations between the severity of higher-order visual impairment and critical visual anatomical networks beyond our assessment of correlation to the PVP. Lastly, investigating differences in visual processing deficits in patients with diffuse and focal white matter injury could provide insight into white matter’s role in remapping abilities and recovery.

## Supplemental Text

### Methods of CVI Cohort

Participants formally diagnosed with CVI (n=17, mean age: 19.1 years, range: 8-31 years; median: 20; IQR: 8) were compared to our childhood stroke and control cohorts. CVI participants had varied suspected causes of CVI including ischemic hypoxic/anoxic injury, periventricular leukomalacia, premature birth complications, and seizure. Members of the CVI cohort were recruited from the Boston area. Details of CVI participant recruitment are described in Manley et al. 2022^21^.

### Childhood stroke survivors have similar VR/ET performance to CVI-diagnosed participants

We compared the VR performance metrics of the childhood stroke cohort to a cohort of formally diagnosed CVI participants (mean age: 19.1 years, range: 8-31 years) (n=17). CVI- diagnosed patients had an increased reaction time compared to stroke patients (p=0.0175; stroke: 2617.76±948.72; CVI: 4084.07±2158.77). In the remaining 4 metrics assessed from the dynamic hallway VR assessment, the childhood stroke and CVI cohorts showed similar impairments (success rate: p=0.296; gaze error: p=0.0513; search area: p=0.133; off-screen percentage: p=0.561) (Supp. Fig. 2). This preliminary analysis indicates that the HOVP abilities of childhood stroke patients align more closely with those of CVI-diagnosed participants than healthy controls.

### Laterality of stroke lesion alters VR outcomes

For participants in the childhood stroke cohort with a supratentorial stroke lesion, we assessed if stroke laterality exacerbated HOVP deficits as measured by the VR dynamic hallway test. No significant differences were found in gaze error and search area between left-sided (n=8), right-sided (n=5), or bilateral stroke lesions (n=3). Patients with right hemisphere lesions showed significantly higher success rates than bilateral stroke patients (right hemisphere stroke: 98.89±2.49; bilateral stroke: 96.67±8.48; Kruskal-Wallis: p=0.047) and faster reaction times (right hemisphere stroke: 1792.16±570.32; left hemisphere stroke: 3305.25±721.06; Kruskal-Wallis: p=0.014) than left hemisphere stroke patients (Supp. Fig. 3). Beyond laterality, there were no significant differences in the VR metrics based upon vascular territories affected - MCA, PCA, ACA, cerebellar or other territories (success rate: p=0.156, reaction time: p=0.486; gaze error: p=0.305; search area: p=0.53; off-screen percentage: p=0.727) (Supp. Fig. 4) or type of stroke– ischemic, hemorrhagic, or venous infarction (success rate: p=0.418, reaction time: p=0.832; gaze error: p=0.973; search area: p=0.348; off-screen percentage: p=0.496) (Supp. Fig. 5).

## Supplementary Figures

**Supplementary Table 1:**
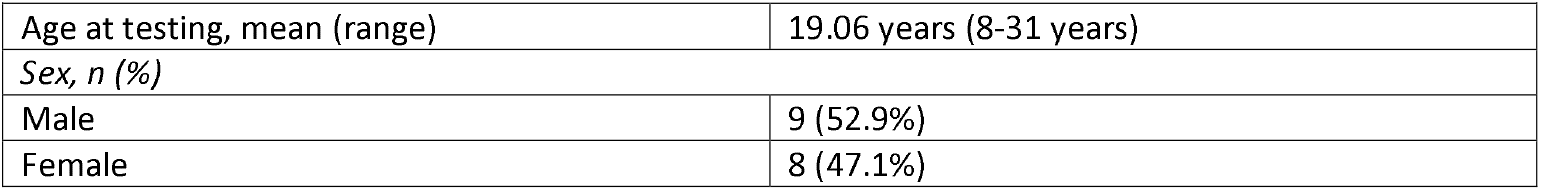
CVI-Diagnosed Cohort Demographics.

**Supplemental Figure 1.**
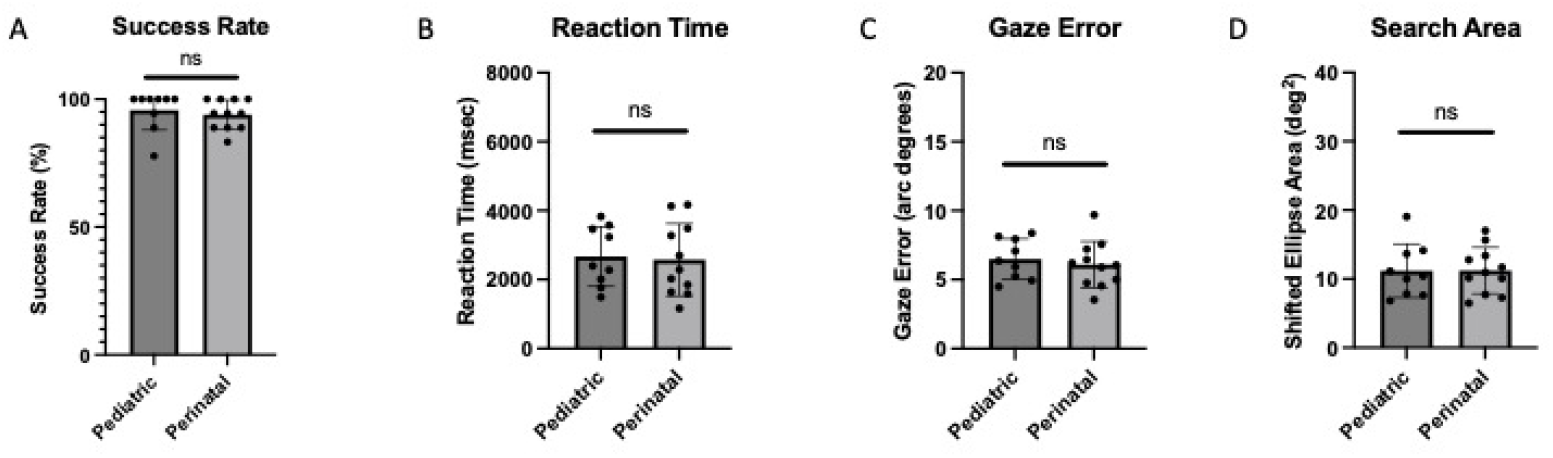
VR performance of pediatric and perinatal stroke patients. Comparing the performance metrics of pediatric and perinatal stroke patients did not show any significant differences across the 4 VR metrics, allowing us to combine these 2 cohorts into one aggregate childhood stroke cohort.

**Supplemental Figure 2.**
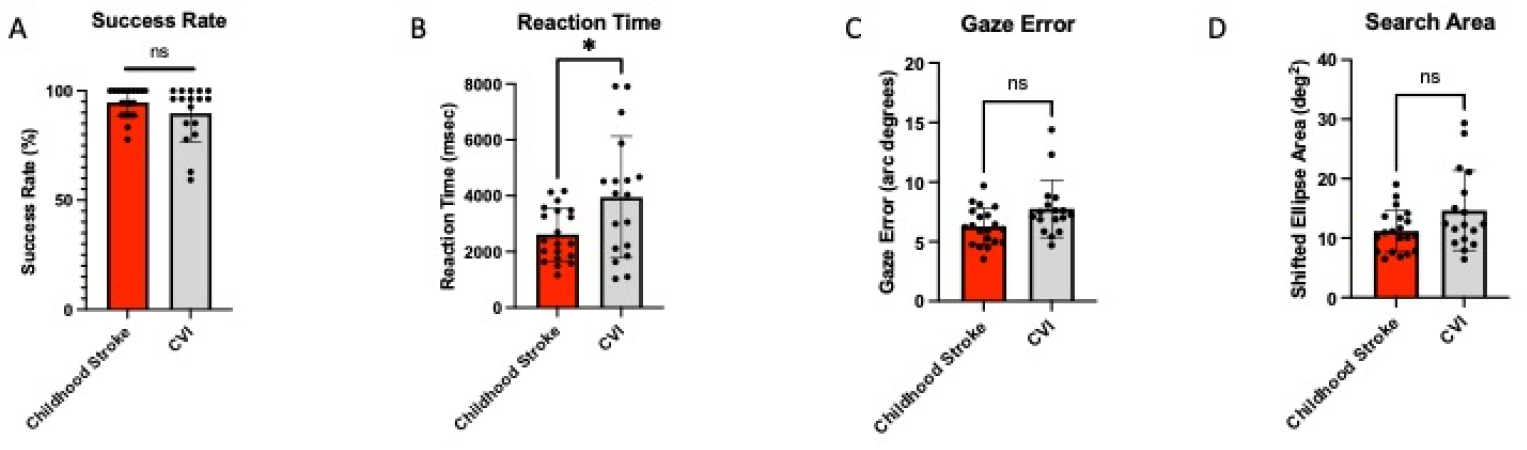
VR task performance of childhood stroke and CVI-diagnosed cohorts. Success rate **(A)**, gaze error **(C)**, and search area **(D)** on the VR search task did not significantly differ between the childhood stroke and CVI-diagnosed cohorts. **(B)** CVI-diagnosed participants exhibited increased reaction time compared to stroke patients (p=0.0175).

**Supplemental Figure 3.**
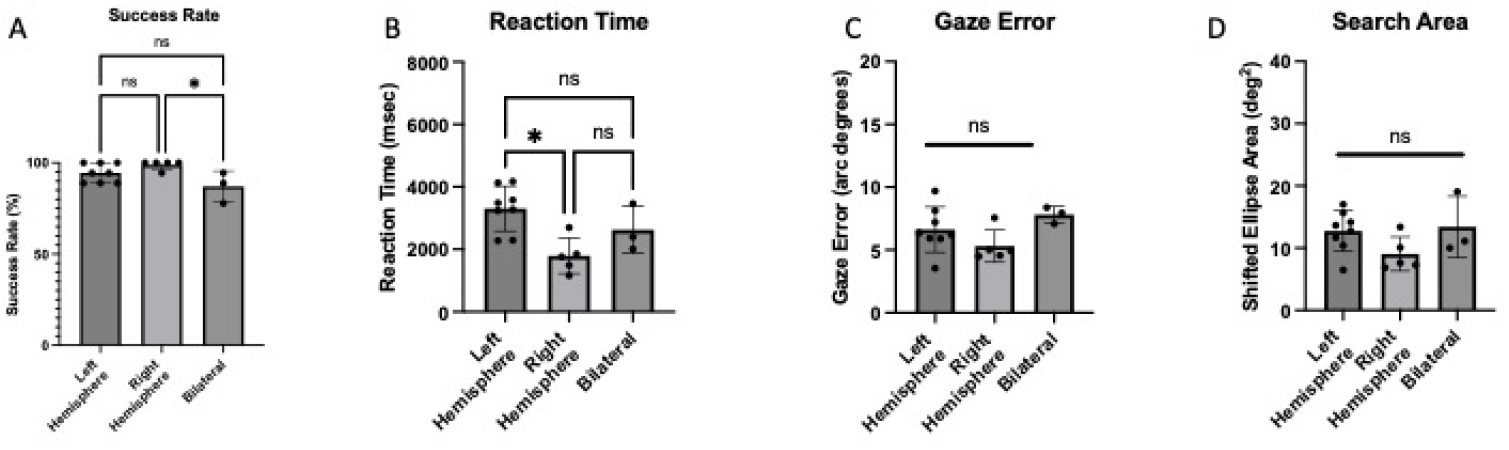
VR task performance of stroke patients as assessed by stroke laterality. **(A)** Childhood stroke patients with right hemispheric injury exhibited significantly increased success rates as compared to stroke patients with bilateral injury (p=0.0469). **(B)** Stroke patients with right hemispheric injury also had significantly faster reaction times than patients with left hemispheric injury (p=0.0137). There were no statistically significant differences in gaze error **(C)** or search area **(D)** on the VR task for patients with left hemispheric, right hemispheric, or bilateral injuries.

**Supplemental Figure 4.**
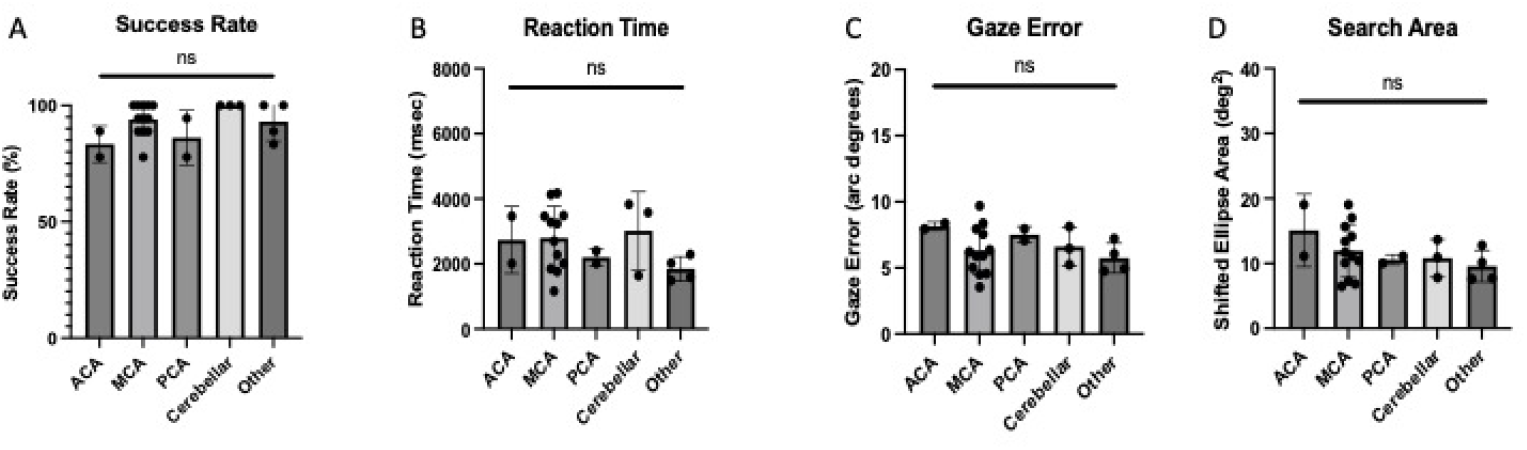
VR task performance of stroke patients by territory affected. Dividing childhood stroke cohort according to the vascular regions affected (ACA, MCA, PCA, cerebellar, or other) did not show any significance across any of the 4 VR metrics captured.

**Supplemental Figure 5.**
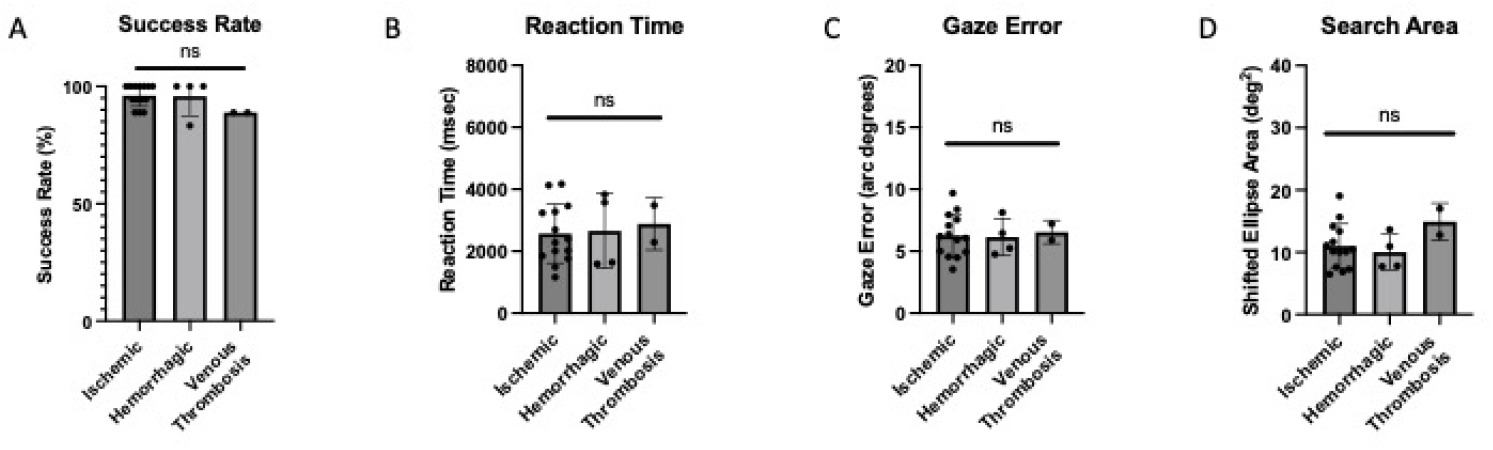
VR task performance of stroke patients by stroke type. Dividing childhood stroke cohort according to their stroke type (ischemic, hemorrhagic, and venous thrombosis) did not show any significance across any of the 4 VR metrics captured.

## Notes

### Competing Interest Statement

The authors have declared no competing interest.

### Funding Statement

This study was funded by National Institute of Health/ National Eye Institute (NIH/NEI R01 EY030973)

### Summary of Updates

Supplemental files updated. Additional analyses to MRI imaging and neuropsychological testing scores assessed. Updating Figures 3 and 4.

## References

1. Lynch JK. Epidemiology and classification of perinatal stroke. Semin Fetal Neonatal Med. Oct 2009;14(5):245–9. doi:10.1016/j.siny.2009.07.001

2. Rawanduzy CA, Earl E, Mayer G, Lucke-Wold B. Pediatric Stroke: A Review of Common Etiologies and Management Strategies. Biomedicines. Dec 20 2022;11(1) doi:10.3390/biomedicines11010002

3. Bernspang B, Asplund K, Eriksson S, Fugl-Meyer AR. Motor and perceptual impairments in acute stroke patients: effects on self-care ability. Stroke. Nov-Dec 1987;18(6):1081–6. doi:10.1161/01.str.18.6.1081

4. Prince M, Lamontagne V, Beauchemin J, et al. Persistent visual perceptual disorders after stroke: Associated factors. British Journal of Occupational Therapy. 2017;80(8):479–485. doi:10.1177/0308022617703240

5. Mercier L, Audet T, Hebert R, Rochette A, Dubois MF. Impact of motor, cognitive, and perceptual disorders on ability to perform activities of daily living after stroke. Stroke. Nov 2001;32(11):2602–8. doi:10.1161/hs1101.098154

6. Rowe F, Brand D, Jackson CA, et al. Visual impairment following stroke: do stroke patients require vision assessment? Age Ageing. Mar 2009;38(2):188–93. doi:10.1093/ageing/afn230

7. Sand KM, Midelfart A, Thomassen L, Melms A, Wilhelm H, Hoff JM. Visual impairment in stroke patients--a review. Acta Neurol Scand Suppl. 2013;(196):52–6. doi:10.1111/ane.12050

8. Westmacott R, MacGregor D, Askalan R, deVeber G. Late emergence of cognitive deficits after unilateral neonatal stroke. Stroke. Jun 2009;40(6):2012–9. doi:10.1161/STROKEAHA.108.533976

9. Lehman LL, Rivkin MJ. Perinatal arterial ischemic stroke: presentation, risk factors, evaluation, and outcome. Pediatr Neurol. Dec 2014;51(6):760–8. doi:10.1016/j.pediatrneurol.2014.07.031

10. Basu AP. Early intervention after perinatal stroke: opportunities and challenges. Dev Med Child Neurol. Jun 2014;56(6):516–21. doi:10.1111/dmcn.12407

11. Bennett CR, Bex PJ, Bauer CM, Merabet LB. The Assessment of Visual Function and Functional Vision. Semin Pediatr Neurol. Oct 2019;31:30–40. doi:10.1016/j.spen.2019.05.006

12. Hart J, Hart J, John. 107Higher-Order Visual Processing. The Neurobiology of Cognition and Behavior. Oxford University Press; 2015:0.

13. Rowe FJ, Hepworth LR, Howard C, Hanna KL, Currie J. Impact of visual impairment following stroke (IVIS study): a prospective clinical profile of central and peripheral visual deficits, eye movement abnormalities and visual perceptual deficits. Disabil Rehabil. Jun 2022;44(13):3139–3153. doi:10.1080/09638288.2020.1859631

14. Rowe F, UK Visg. Visual perceptual consequences of stroke. Strabismus. Jan-Mar 2009;17(1):24–8. doi:10.1080/09273970802678537

15. Rowe F, UK Visg. Symptoms of stroke-related visual impairment. Strabismus. Jun 2013;21(2):150–4. doi:10.3109/09273972.2013.786742

16. Hepworth L, Rowe F, Walker M, et al. Post-stroke Visual Impairment: A Systematic Literature Review of Types and Recovery of Visual Conditions. Ophthalmology Research: An International Journal. 01/10 2016;5:1–43. doi:10.9734/OR/2016/21767

17. Aravind G, Lamontagne A. Effect of visuospatial neglect on spatial navigation and heading after stroke. Ann Phys Rehabil Med. Jul 2018;61(4):197–206. doi:10.1016/j.rehab.2017.05.002

18. Painter DR, Norwood MF, Marsh CH, et al. Immersive virtual reality gameplay detects visuospatial atypicality, including unilateral spatial neglect, following brain injury: a pilot study. J Neuroeng Rehabil. Nov 23 2023;20(1):161. doi:10.1186/s12984-023-01283-9

19. Rowe F, Wright D, Brand D, et al. Reading difficulty after stroke: ocular and non ocular causes. Int J Stroke. Oct 2011;6(5):404–11. doi:10.1111/j.1747-4949.2011.00583.x

20. Vancleef K, Colwell MJ, Hewitt O, Demeyere N. Current practice and challenges in screening for visual perception deficits after stroke: a qualitative study. Disabil Rehabil. May 2022;44(10):2063–2072. doi:10.1080/09638288.2020.1824245

21. Manley CE, Bennett CR, Merabet LB. Assessing Higher-Order Visual Processing in Cerebral Visual Impairment Using Naturalistic Virtual-Reality-Based Visual Search Tasks. Children (Basel). Jul 26 2022;9(8) doi:10.3390/children9081114

22. Manley CE, Walter K, Micheletti S, et al. Object identification in cerebral visual impairment characterized by gaze behavior and image saliency analysis. Brain Dev. Sep 2023;45(8):432–444. doi:10.1016/j.braindev.2023.05.001

23. Corre CS, Bambery M, Bennett CR, et al. Characterizing visual processing deficits in cerebral adrenoleukodystrophy. Brain Dev. Oct 12 2024; doi:10.1016/j.braindev.2024.09.008

24. Merabet LB, Mayer DL, Bauer CM, Wright D, Kran BS. Disentangling How the Brain is “Wired” in Cortical (Cerebral) Visual Impairment. Semin Pediatr Neurol. May 2017;24(2):83–91. doi:10.1016/j.spen.2017.04.005

25. Martin MB, Santos-Lozano A, Martin-Hernandez J, et al. Cerebral versus Ocular Visual Impairment: The Impact on Developmental Neuroplasticity. Front Psychol. 2016;7:1958. doi:10.3389/fpsyg.2016.01958

26. Chang MY, Merabet LB, Group CVIW. Special Commentary: Cerebral/Cortical Visual Impairment Working Definition: A Report from the National Institutes of Health CVI Workshop. Ophthalmology. Dec 2024;131(12):1359–1365. doi:10.1016/j.ophtha.2024.09.017

27. Boonstra FN, Bosch DGM, Geldof CJA, Stellingwerf C, Porro G. The Multidisciplinary Guidelines for Diagnosis and Referral in Cerebral Visual Impairment. Front Hum Neurosci. 2022;16:727565. doi:10.3389/fnhum.2022.727565

28. Morelli F, Aprile G, Martolini C, et al. Visual Function and Neuropsychological Profile in Children with Cerebral Visual Impairment. Children (Basel). Jun 19 2022;9(6) doi:10.3390/children9060921

29. Crawford LB, Golomb MR. Childhood Stroke and Vision: A Review of the Literature. Pediatr Neurol. Apr 2018;81:6–13. doi:10.1016/j.pediatrneurol.2017.11.007

30. Luckman J, Chokron S, Michowiz S, et al. The Need to Look for Visual Deficit After Stroke in Children. Front Neurol. 2020;11:617. doi:10.3389/fneur.2020.00617

31. McDowell N, Budd J. The Perspectives of Teachers and Paraeducators on the Relationship between Classroom Clutter and Learning Experiences for Students with Cerebral Visual Impairment. Journal of Visual Impairment & Blindness. 2018;112(3):248–260. doi:10.1177/0145482x1811200304

32. Dutton GN, McKillop EC, Saidkasimova S. Visual problems as a result of brain damage in children. Br J Ophthalmol. Aug 2006;90(8):932–3. doi:10.1136/bjo.2006.095349

33. Sabel BA, Henrich-Noack P, Fedorov A, Gall C. Vision restoration after brain and retina damage: the “residual vision activation theory”. Prog Brain Res. 2011;192:199–262. doi:10.1016/B978-0-444-53355-5.00013-0

34. Wechsler D. Wechsler intelligence scale for children–Fifth Edition (WISC-V). Bloomington, MN: Pearson. 2014;

35. Malone LA, Felling RJ. Pediatric Stroke: Unique Implications of the Immature Brain on Injury and Recovery. Pediatr Neurol. Jan 2020;102:3–9. doi:10.1016/j.pediatrneurol.2019.06.016

36. Berman JI, Glass HC, Miller SP, et al. Quantitative Fiber Tracking Analysis of the Optic Radiation Correlated with Visual Performance in Premature Newborns. American Journal of Neuroradiology. 2009;30(1):120–124. doi:10.3174/ajnr.A1304

37. Bassi L, Ricci D, Volzone A, et al. Probabilistic diffusion tractography of the optic radiations and visual function in preterm infants at term equivalent age. Brain. Feb 2008;131(Pt 2):573–82. doi:10.1093/brain/awm327

38. Herbet G, Zemmoura I, Duffau H. Functional Anatomy of the Inferior Longitudinal Fasciculus: From Historical Reports to Current Hypotheses. Front Neuroanat. 2018;12:77. doi:10.3389/fnana.2018.00077

39. Tavor I, Yablonski M, Mezer A, Rom S, Assaf Y, Yovel G. Separate parts of occipito-temporal white matter fibers are associated with recognition of faces and places. Neuroimage. Feb 1 2014;86:123–30. doi:10.1016/j.neuroimage.2013.07.085

40. Benson DF, Segarra J, Albert ML. Visual Agnosia-Prosopagnosia: A Clinicopathologic Correlation. Archives of Neurology. 1974;30(4):307–310. doi:10.1001/archneur.1974.00490340035007

41. Kim S-H, Jeon H-E, Park C-H. Relationship between Visual Perception and Microstructural Change of the Superior Longitudinal Fasciculus in Patients with Brain Injury in the Right Hemisphere: A Preliminary Diffusion Tensor Tractography Study. Diagnostics. 2020;10(9):641.

42. Mishkin M, Ungerleider LG, Macko KA. Object vision and spatial vision: two cortical pathways. Trends in Neurosciences. 1983;6:414–417. doi:10.1016/0166-2236(83)90190-X

43. Haxby JV, Grady CL, Horwitz B, et al. Dissociation of object and spatial visual processing pathways in human extrastriate cortex. Proc Natl Acad Sci U S A. Mar 1 1991;88(5):1621–5. doi:10.1073/pnas.88.5.1621

44. Bennett CR, Bauer CM, Bailin ES, Merabet LB. Neuroplasticity in cerebral visual impairment (CVI): Assessing functional vision and the neurophysiological correlates of dorsal stream dysfunction. Neurosci Biobehav Rev. Jan 2020;108:171–181. doi:10.1016/j.neubiorev.2019.10.011

45. Lennartsson F, Ohnell H, Jacobson L, Nilsson M. Pre- and Postnatal Damage to the Retro-Geniculate Visual Pathways Cause Retinal Degeneration Predictive for Visual Function. Front Hum Neurosci. 2021;15:734193. doi:10.3389/fnhum.2021.734193

46. Koenraads Y, Porro GL, Braun KPJ, Groenendaal F, de Vries LS, van der Aa NE. Prediction of visual field defects in newborn infants with perinatal arterial ischemic stroke using early MRI and DTI-based tractography of the optic radiation. Eur J Paediatr Neurol. Mar 2016;20(2):309–318. doi:10.1016/j.ejpn.2015.11.010

47. Hoyt CS. Visual function in the brain-damaged child. Eye (Lond). Apr 2003;17(3):369–84. doi:10.1038/sj.eye.6700364

