## Supplemental Data for "Involvement of the Posterior Visual Pathway Correlates with Higher-Order Visual Impairment in Childhood Stroke Patients detected by Virtual Reality/Eye Tracking Paradigm"

**Supplementary Table and Figures:**

| **Supplementary Table 1: CVI-Diagnosed Cohort Demographics** | |
| --- | --- |
| Age at testing, mean (range) | 19.06 years (8-31 years) |
| *Sex, n (%)* | |
| Male | 9 (52.9%) |
| Female | 8 (47.1%) |

**Supplemental Figure 1. VR performance of pediatric and perinatal stroke patients.** Comparing the performance metrics of pediatric and perinatal stroke patients did not show any significant differences across the 4 VR metrics, allowing us to combine these 2 cohorts into one aggregate childhood stroke cohort.

**Supplemental Figure 2. VR task performance of childhood stroke and CVI-diagnosed cohorts.** Success rate (**A**), gaze error (**C**), and search area (**D**) on the VR search task did not significantly differ between the childhood stroke and CVI-diagnosed cohorts. (**B**) CVI-diagnosed participants exhibited increased reaction time compared to stroke patients (p=0.0175).

**Supplemental Figure 3. VR task performance of stroke patients as assessed by stroke laterality.** (**A**) Childhood stroke patients with right hemispheric injury exhibited significantly increased success rates as compared to stroke patients with bilateral injury (p=0.0469). (**B**) Stroke patients with right hemispheric injury also had significantly faster reaction times than patients with left hemispheric injury (p=0.0137). There were no statistically significant differences in gaze error (**C**) or search area (**D**) on the VR task for patients with left hemispheric, right hemispheric, or bilateral injuries.

**Supplemental Figure 4. VR task performance of stroke patients by territory affected.** Dividing childhood stroke cohort according to the vascular regions affected (ACA, MCA, PCA, cerebellar, or other) did not show any significance across any of the 4 VR metrics captured.

**Supplemental Figure 5. VR task performance of stroke patients by stroke type.** Dividing childhood stroke cohort according to their stroke type (ischemic, hemorrhagic, and venous thrombosis) did not show any significance across any of the 4 VR metrics captured.
